# Adjusting COVID-19 Reports for Countries’ Age Disparities: A Comparative Framework for Reporting Performances

**DOI:** 10.1101/2020.08.31.20185223

**Authors:** Enes Eryarsoy, Dursun Delen, Behrooz Davazdahemami

## Abstract

**Objectives:** The COVID-19 outbreak has impacted distinct health care systems differently. While the rate of disease for COVID-19 is highly age-variant, there is no unified and age/gender-inclusive reporting taking place. This renders the comparison of individual countries based on their corresponding metrics, such as CFR difficult. In this paper, we examine cross-country differences, in terms of the age distribution of symptomatic cases, hospitalizations, intensive care unit (ICU) cases, and fatalities. In addition, we propose a new quality measure (called dissonance ratio) to facilitate comparison of countries’ performance in testing and reporting COVID-19 cases (i.e., their reporting quality).

**Methods:** By combining population pyramids with estimated COVID-19 age-dependent conditional probabilities, we bridge country-level incidence data gathered from different countries and attribute the variability in data to country demographics.

**Results:** We show that age-adjustment can account for as much as *a 22-fold difference* in the expected number of fatalities across different countries. We provide case, hospitalization, ICU, and fatality breakdown estimates for a comprehensive list of countries. Also, a comparison is conducted between countries in terms of their performance in reporting COVID-19 cases and fatalities.

**Conclusions:** Our research sheds light on the importance of and propose a methodology to use countries’ population pyramids for obtaining accurate estimates of the healthcare system requirements based on the experience of other, already affected, countries at the time of pandemics.

## Introduction

The first COVID-19 outbreak took place in the city of Wuhan in the Hubei province of China. Despite strict and robust prevention measures taken in the city, the virus has spread the rest of the world in a matter of a few weeks. Within three months, the World Health Organization (WHO) declared the outbreak a pandemic. While some more significantly than others, the virus has taken its toll on all countries with no exception. The global impact of COVID-19 has been very profound and probably unprecedented since the Spanish flu (H1N1 influenza circa 1918). Due to the novelty of the virus, and the nonexistence of vaccination, health professionals have been trying to cope with the pandemic using symptomatic treatment regimes.

Many governments are seeking out forming different strategies that involve mitigating the spread until a method of prevention or a well-defined, and a successful treatment regime is found (1). The main focus of such mitigation effects is to alleviate the burden on healthcare systems by spreading out the diffusion of cases over a more extended period of time. While trying to achieve this, governments also face many uncertainties. One such uncertainty involves the absence of proven methods to accurately estimate the potential demand for healthcare services (2,3).

At the time of this paper’s writing, several governments, such as Italy and Spain, already had over 100% health services capacity utilization, while others were about to experience a similar influx of critical patients. It is clear that governments are in need of better understanding the dynamics of the spread for optimal or near-optimal resource allocation decisions. Unfortunately, due to the emergency and the gravity of the pandemic, and the lack of scantily found hard evidence causes, such decisions have been made through the seat-of-the-pants approaches.

Perhaps one of the reasons behind the lack of evidence is that there is no obvious way to map reports and studies pertaining to one country into another. Many regional differences make this mapping and transfer of the learnings and knowledge over to another domain particularly difficult. For the case of COVID-19, gender and age of the patient populations seem to be among the key drivers of such differences. Academics are acting swiftly to enrich the medical literature by reporting their findings on the virus-related population characteristics, diffusion patterns, treatment regimes, case dynamics, hospitalizations, ICU usages, and fatalities(4-7).

In this paper, as the first objective, we build on the studies and reports that involve age-based clinical fatality risks (CFR), infection fatality risks (IFR), hospitalizations, ICU usages, and fatal outcomes. Using the latest literature as well as expert opinions, we attempt to combine data from different regions in order to estimate and highlight: (i) country-level differences, and (ii) healthcare system demands for individual age groups. Specifically, using the data from six different countries, we study the spread of the virus for different age groups.

It is obvious that different countries, either due to their different policies or because of different healthcare infrastructures, do not perform equally well in conducting diagnostic tests and reporting a number of cases. Due to this fact, even countries with similar demographics and social distancing policies may report highly inconsistent numbers. Such inconsistencies will then make studying the disease’s epidemiological characteristics challenging.

As the second objective, this paper seeks to propose an approach to compare the reporting performance of countries during the COVID-19 pandemic. For this purpose, we rely on the countries’ similarities in terms of their age pyramids as well as the stage of the disease and assume that similar countries should experience similar population- and age-standardized numbers for their infections and death tolls. Considering US as the baseline, we then calculate a dissonance ratio for each country as a standardized measure of their performance in reporting cases and mortalities (i.e., a measure of the quality of their reports).

The rest of this paper is organized as follows: in the next section, a review of relevant studies from literature is provided. Following that, we describe our method and elaborate on the data used. The final section is dedicated to the findings and related discussions.

## Background

The rapid spread of the novel coronavirus has even made the calculation of the rate of spread difficult. One frequently used way of measuring the spread is by computing the average number of secondary cases, or infections, that each case generates. This is known as the R-naught (*R*_0_) (a.k.a., reproduction number) of the virus. The *R*_0_*’s* time- and place-dependent nature (typically smaller in the South Asian countries, depending on social distancing interventions such as case isolation, partial or complete lockdowns, school closure, and distance working by the local authorities) is making modeling the spread of the virus a moving target (8).

Even though the literature on COVID-19 is rapidly expanding, there is still a lack of consensus among academics and other scientists on the dynamics of the spread. Different estimates, for instance, are reported for the disease’s *R*_0_ ranging from 1.94 through 6.7 (6,9,10). This can perhaps be attributed to many reasons, such as the unpreparedness to a pandemic at this level, the lack of unified reporting systems due to diversity of health systems across the world, and the novelty of the pandemic itself.

Studies also report varying figures for other epidemiological measures such as CFR or IFR. Several underlying reasons may explain these variabilities. Perhaps one of the most plausible reasons is the abundance of undocumented cases. In their study, Li et al. (10) highlight that one of the reasons for the rapid spread of the virus is lack of documentation. They estimate that around 86% of all infections were undocumented. Another study from South Korea suggests similar undocumented case percentages at around 55-86% (11). News also suggests that even mortality cases often go unreported. A recent article in The Economist (12) highlighted stark differences between the number of expected death cases (including those attributed to COVID-19) and the actual death cases. Their estimation, based on regions’ normal death rates, suggests the actual death-toll of the novel COVID-19 being more than double of what is being reported in different regions in Italy, Spain, and France. Perhaps this may be one of the reasons for conflicting CFR and IFR figures reported in the literature. While some studies suggested an estimated case fatality risk of as high as 7.2% (13) in Italy, other studies reported a CFR of 3.4% in China (14), and 2.3% using the age-adjusted Diamond Princess cruise ship data (15). More recent works report somewhat lower case fatality rates circa 1.4-1.5%% (16,17), both using data from Wuhan.

Similar variations also hold for IFR. Studies report IFRs as low as 0.5% using the Diamond Princess cruise ship data (15), 0.94%, and 0.657% using Wuhan data (18) and (19), respectively. Even though these numbers significantly differ from each other, there seems to be (i) convergence to a CFR of 1.5% over time, (ii) CFR/IFR ratio appears to hover around 2-3, indicating as much as 40-70% asymptomatic cases of the virus.

One of the apparent reasons behind the differences in reported CFRs and IFRs is the different demographics in different countries. As the virus affects the elderly more than the young, the virus takes a different toll on each country, depending on its age demographics (15,20). There may be several other reasons, including the fact that the age distribution of the infected population may differ from the overall age distribution. Hence, *R*_0_ may significantly differ across age groups. However, there is a lack of by-age (and gender)-group reporting. For instance, Raj Bhopal (21), in his article expresses the importance and urgency of adjusting/reporting age and gender breakdowns of COVID-19 related data.

In this paper we pursue two objectives; First, we take into account the cross-country age disparities to handle such inconsistencies by providing country-independent conditional probabilities for each age group and then estimating the number of infections, hospitalizations, ICU visits, and mortality for each country given its age distribution. To this end, we assume that deaths, hospitalizations, and ICU usages are proxy measures for COVID-19 spread, also that similar spread patterns apply to each age group across countries—the virus is identical across all countries. We also demonstrate some evidence for the acceptability of these assumptions.

Second, relying on our findings from the first part, we propose a measure to assess the quality of testing and reporting COVID-19 cases across countries and comparing countries’ performance in those manners. We believe that lack of such a measure in the literature has led to many studies relying on poor quality COVID-19 data in their analyses, which may lead to misleading conclusions.

### Method and Dataset

Many of the existing studies that are focused on modeling the spread of the COVID-19 have been using a few different models such as susceptible-infected-recovered (SIR) and its covariates (mainly SEIR: susceptible-exposed-infected-recovered, or Sidarthe model) (22-26). These studies often ignore age-dependent variations from one country to another, or are limited to one country, or sometimes two (1).

In this study, instead of computing the spread of the virus, we look at the results of different scenarios. Specifically, we design three different spread scenarios (mild, moderate, and severe) by transferring knowledge learned from the seasonal flu pandemic as well as existing estimates for COVID-19 spread measures (R_0_, proportion of symptomatic infections (C), and proportion of reported cases) from the literature. Then taking into account expert opinions, we choose the best scenario (closest one to their opinions).

The best scenario is then used, along with the US (our baseline country) population age distribution, to calculate a set of conditional probabilities to estimate probability of infection, hospitalization, ICU admission, and death for a given person from each age group. At the end, the calculated probabilities are applied to the countries’ age pyramids to estimate cases, hospitalizations, ICU usage, and fatalities in each country (adjusted for their population age distribution).

### Scenarios and Expert opinions

While aggressive quarantines and enforcing/recommending social distancing can change the outcome of the burden on healthcare systems, the primary health care capacities are the bottleneck for almost all countries. The size of the susceptible population typically depends on different *R*_0_ values. By enforcing/recommending social distancing, governments attempt to mitigate the situation and reduce this number (*Figure 1*).

**Figure 1.**
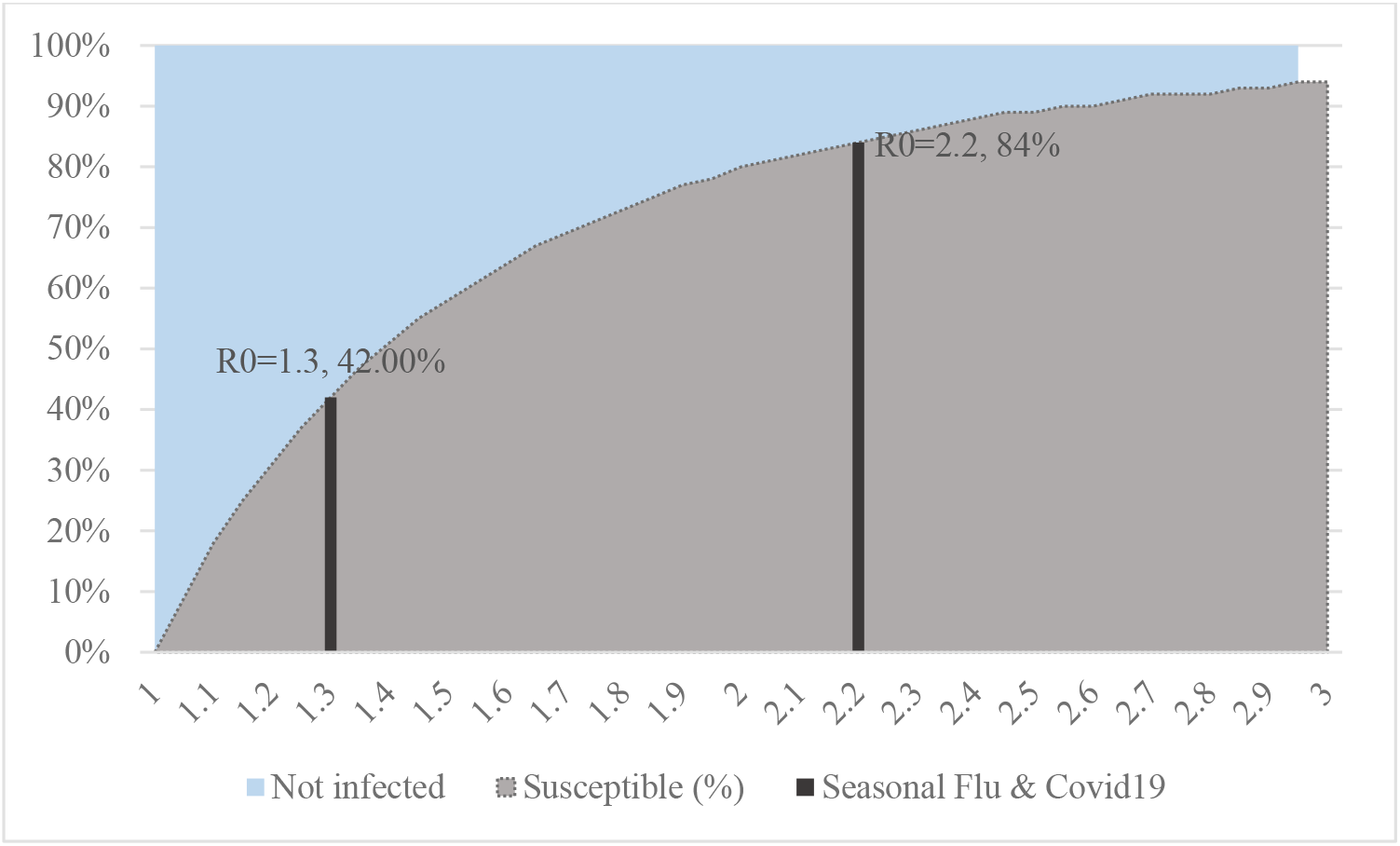
Different *R*_0_ values and corresponding estimated percent of susceptible populations.

The novel coronavirus is often compared and contrasted against seasonal flu. Using CDC numbers over the last two flu seasons (2017-2019), we estimate the following table for the seasonal flu for comparison purposes.

**Table 1.** The seasonal flu numbers from the CDC.

| Seasonal flu (based on CDC data) | US Cases | As % of Susceptible Population | Per 1M |
| --- | --- | --- | --- |
| Susceptible population ($R_0=1.3$) | 134.4M | 100.00% | 420,000 |
| Population with symptoms | 40M | 29.80% | 121K |
| Medical Visits | 15M | 11.10% | 45.5K |
| Hospitalization | 0.6M | 0.44% | 1,823 |
| Fatality | 40K | 0.03% | 121 |
| CFR | 0.03% |  |  |

We construct an analogous table to seasonal flu using expert opinions (27,28) (See Table 2). To this end, we constructed different scenarios severe (Scenario 1) to mild (Scenario 3) based on the estimates provided by the existing literature. We construct this table for the United States. By making use of suggested estimates based on (27,29), we create a range of possible *R*_0_ values (i.e., 1.5-2.2) to estimate the percentage of susceptible population. Using the literature, we then estimate upper and lower limits for symptomatic cases, reported cases (not all of the symptomatic cases are reported), hospitalizations (as a percentage of reported cases), as well as ICU cases (in terms of cases), and fatalities in an age-adjusted manner.

**Table 2.** Different COVID-19 spread estimates for the United States (numbers are in millions)

| COVID-19 | Scen. 1 | Scen. 2 | Scen. 3 | Expert Opinion 1 | Expert Opinion 2 |
| --- | --- | --- | --- | --- | --- |
| Susceptible population (overall $R_0 = \{2.2^1, 1.8, 1.5^2\}$) | 276.4 | 240.2 | 190.8 | 160-210M | - |
| The population with symptoms $\{.50^3, .35, .20^4\}$(C) | 138 | 84 | 38 | - | - |
| Reported cases (as the ratio of symptomatic cases) $\{.11^5, .15, .20^6\}$ | 27.6 | 13 | 4.2 | - | - |
| Hospitalizations (36%<sup>7,8</sup> of reported cases)(H) | 10 | 4.5 | 1.5 | 2.4-21M | - |
| ICU patients (% of Hospitalizations–7.4%<sup>9,10</sup>)(I) | 0.7 | 0.34 | 0.11 | - | ~0.08<sup>11</sup> |
| Fatalities (D) | 0.41 | 0.19 | 0.063 | 0.2-1.7M | 0.03-0.12 |
| CFR $\{1.5^{12,13}\}$ | 1.50% | 1.50% | 1.50% | | |

<sup>1</sup> (10)**Error! Bookmark not defined.** : estimates $R_0$ value as 2.2 for Wuhan data.
<sup>2</sup> (30) estimates $R_0$ value as 1.5 for South Korea where the virus is relatively better contained
<sup>3</sup> (17) finds a 50% chance of developing symptoms using Wuhan data.
<sup>4</sup> (31) suggests that the ratio of asymptomatic cases could be as high as 80%.
<sup>5</sup> (32) uses South Korean data and similar age corrections and estimate that only around 11% of cases are reported in the US.
<sup>6</sup> (33) uses an estimation interval of 40-60% in their analysis. However, we construct our scenarios using similar to the US-based study.
<sup>7</sup> (34) CDC estimates also yield a similar number in the US. However, the data has no age breakdown and includes some unknown cases.
<sup>8</sup> The rate is estimated as 0.36 using conditional probabilities and age distributions based on Spanish Data (35)
<sup>9</sup> (29) estimates that around a fifth of hospitalization days will require ICU stays for the US. However, they do not provide projections based on the number of hospitalization and ICU cases. Their numbers are in seem consistent with Spanish data (35).
<sup>10</sup> The crude rate is estimated as 7.4% of total hospitalizations using Spanish Data (35)**Error! Bookmark not defined.** . However, each case stays in ICU for an average of 15 days (36). ( $0.074 \times 15 = 1.11$ )
<sup>11</sup> Assuming a 10-day average duration of stay.
<sup>12</sup>(16,17) indicated a CFR of 1.5%.
<sup>13</sup> (37) indicates 0.7% CFR in Germany; such numbers usually come from countries where the spread is better contained.

Estimating fatalities is challenging since case fatality rates depend on a number of factors, including:

i. The number of tests (and therefore, the number of positive cases) conducted each individual country. Many countries—excluding countries such as Iceland, where a significant portion of the population was tested—conduct selective testing. This may involve a selection bias where only the people with severe enough symptoms may be tested.
ii. The delay between the symptom onsets and the time of deaths.
iii. The varying levels of adequacy/inadequacy of the healthcare systems.
iv. The rates of smoking or the prevalence of chronical illnesses. We chose to use CFR of 1.5% for the United States for our analysis.

Based on the characteristics of the COVID-19, the Center for Disease Control (CDC) estimated that 2.4 to 21 million Americans would require hospitalization, and a death-toll of as much as 480,000 may be expected (27). According to the same projection, the death toll could be any figure from 200,000 to as high as 1.7 million. Another more recent estimate assuming full social distancing through May (as of April 8, 2020), the White House estimated this figure to fall between 30,000 and 126,000 (28). These numbers are shown in the last two columns of Table 2. Given the closeness of estimates made by Scenario 3 (the mildest scenario) and the expert opinions, we rely on that scenario for establishing the conditional probabilities discussed in the following section.

### Conditional Probabilities

In this study, we use countries’ age distributions (population pyramids) and data involving different countries to create country-independent conditional probabilities for each age group. The severity of COVID-19 is also gender-dependent. However, due to the unavailability of data, we did not take gender into account.

As discussed earlier, we report our findings using the mildest of the three scenarios since its estimates turned out to be more consistent with the expert opinions (see Table 2). We use the following notation to formulate the conditional probabilities for each age group:

E: Events, E = {C: Case, A: Age, H: Hospitalization, I: Intensive Unit Care, D: Death)
*P*(*C*): The probability of being infected with symptoms. Using the scenario-2 with
*R*_0_=1.8, we estimate the proportion of the susceptible population as 0.73, with a 35% probability of developing symptoms: *P*(*C*) *=* 0.35 × 0.73
*P*(*R*): The probability of being a reported case: *P*(*R*) *= P*(*C*) *×*.15
*P*(*H*): The probability of hospitalization. This number depends on the percentage of reported cases, as well as the size of the population with symptoms. Using 0.15 and 36% of the rate of hospitalization we use (*P*(*H*) *= P*(*C*) *×*.11 ×.36)
*P*(*I*): The probability of needing ICU (*P*(*I*) *= P*(*H*) × 0.074)
*P*(*D*): The probability of death forthe cases (CFR) (*P*(*D*) *= P*(*R*) *×*.015)
*P*(*A_i_*): The probability of each age group *i* for a given country (using the country population pyramid)
*P*(*C|A_i_*): The probability of being infected with symptoms given age group *i*

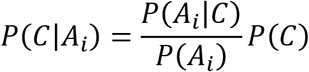

where *P*(*A_i_|C*): The probability of the age group *i*, given case.

We compute other conditional probabilities similarly for events {H, I, D} and then use them to simulate the mild scenario breakdowns for the United States. By using population pyramids and the US data, we replicate the same scenario for each individual country and report the results (per 1 million residents).

### Assessment of Quality of COVID-19 Case Reporting

The second question this study seeks to address is that during the COVID-19 pandemic, how well different affected countries performed in conducting tests and reporting the cases. In order to answer this question, it is critical to have a base for comparing the countries’ performance with one another (i.e., to assess their relative quality of reporting). For this purpose, through a series of calculations described below, we estimate an age-standardized expected number of cases and fatalities for each country, such that countries can be compared regardless of their age distributions. The procedure is explained below.

Using conditional probabilities, we calculate a set of successive measure for each country described below:

a. Age-Adjustment Fatality Coefficient (AAFC): We calculate the age-adjusted expected number of deaths by setting the global fatality baseline coefficient at “1” for the world. The coefficient value higher than “1” typically corresponds to an aging population. For instance, we estimate AAFC for Japan (one of the oldest nations in the world) as 3.41 (1037/204 from Table 1) and AAFC for Niger (the youngest nation) as 0.15. A 22-fold difference.
b. Age-Adjusted ICU Coefficient (AAICUC): The number of expected ICU cases corresponding to corresponding to AAFC of one. Calculated using the formula:

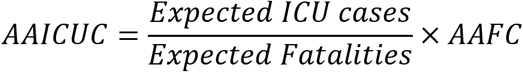 Countries with AAICUC of greater than the global average, 2.26, typically are expected to have heavier demand per capita for their intensive care units.
c. Age-Adjusted Hospitalization Coefficient (AAHC): The number of expected hospitalization cases corresponding to AAFC of one. Calculated using the formula:

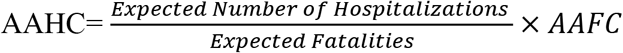
d. Cumulative Fatalities (CF): Number of cumulative fatalities for each country as of May 13.
e. Day-Adjusted Cumulative Cases (DACC): Using a sliding lag window, we estimated the average number of days from the case reported to recovery (or fatality) as 8 days (38). Therefore, DACC corresponds to the reported cumulative number of cases 8 days prior to CF numbers.
f. Actual Case Fatality Risk (ACFR): Calculated as CF/DACC
g. Expected Infections per Fatality (EIPF): A recent study estimated the IFR of COVID-19 to be 1.3 for the US (39). Using this value for the US by adjusting AAFC of the US yields:

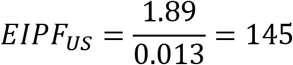 The figure represents the number of infections in the US, corresponding to one fatality at the baseline. Values for other countries are calculated using

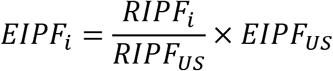 Countries with higher numbers are expected to report more cases than that of the US.
h. Reported Infections per Fatality (RIPF): Calculated as 1/ACFR for each country.
i. Age-adjusted Reported Fatalities per 1M (AARFPM): Some countries, given their population pyramids, are expected to report more cases. This column age and population (per 1M) adjusts the CF values for each country. Note that AARFPM values depend on the stage of the spread.
j. Dissonance Ratio (DR): Calculated as RIPF/ACFR. A value (typically less than 1) that indicates how well two or more countries with roughly the same AARFPM values performed in testing and reporting COVID-19 cases. Smaller DR values indicate possible under-reporting by the corresponding country.

## Results

### Age-adjusted Estimates of COVID-19

Unfortunately, the only source of data that explicitly provided the age breakdowns of hospitalization and ICU cases (P(A|H), and P(A|I)) we could found was Spain (35). After comparing the estimated P(H) and P(I)s from Spain data and the data from the Institute for Health Metrics and Evaluation (28), we concluded the numbers are consistent and decided to use age breakdowns from the Spanish dataset as the baseline for calculating conditional probabilities. A sample table, including some of the probabilities using the reports by the Spanish Ministry of Health, is given in Table 3.

**Table 3.** Probabilities and conditional probabilities for Spain

| Age Group | $P(A)$ | #Inf. Cases | $P(A\vert C)$ | #Hosp. | $P(A\vert H)$ | #ICU cases | $P(A\vert I)$ | #Deaths | $P(A\vert D)$ |
| --- | --- | --- | --- | --- | --- | --- | --- | --- | --- |
| **0-9** | 9.3% | 130 | 0.6% | 35 | 0.45% | 1 | 0% | 0 | 0.0% |
| **10-19** | 10.0% | 226 | 1.1% | 20 | 0.26% | 1 | 0% | 1 | 0.1% |
| **20-29** | 10.0% | 1,352 | 6.6% | 200 | 2.6% | 10 | 2% | 4 | 0.5% |
| **30-39** | 13.2% | 2,386 | 11.7% | 431 | 5.6% | 18 | 3% | 3 | 0.4% |
| **40-49** | 17.0% | 3,190 | 15.6% | 778 | 10.1% | 45 | 8% | 9 | 1.1% |
| **50-59** | 14.9% | 3,433 | 16.8% | 1,074 | 13.9% | 106 | 18% | 20 | 2.5% |
| **60-69** | 11.1% | 3,179 | 15.6% | 1,432 | 18.6% | 162 | 28% | 63 | 7.8% |
| **70-79** | 8.4% | 3,304 | 16.1% | 1,858 | 24.1% | 192 | 34% | 164 | 20.4% |
| **80+** | 6.2% | 3,271 | 16.0% | 1,871 | 24.3% | 38 | 7% | 541 | 67.2% |

Using age-corrections via conditional probabilities also shows that reported numbers are quite consistent across-countries (Table 4). As shown in this table, there is a 0.97 correlation between the estimated and the reported age group probabilities for the United States, suggesting the acceptable performance of the proposed approach for age adjustments.

**Table 4.**
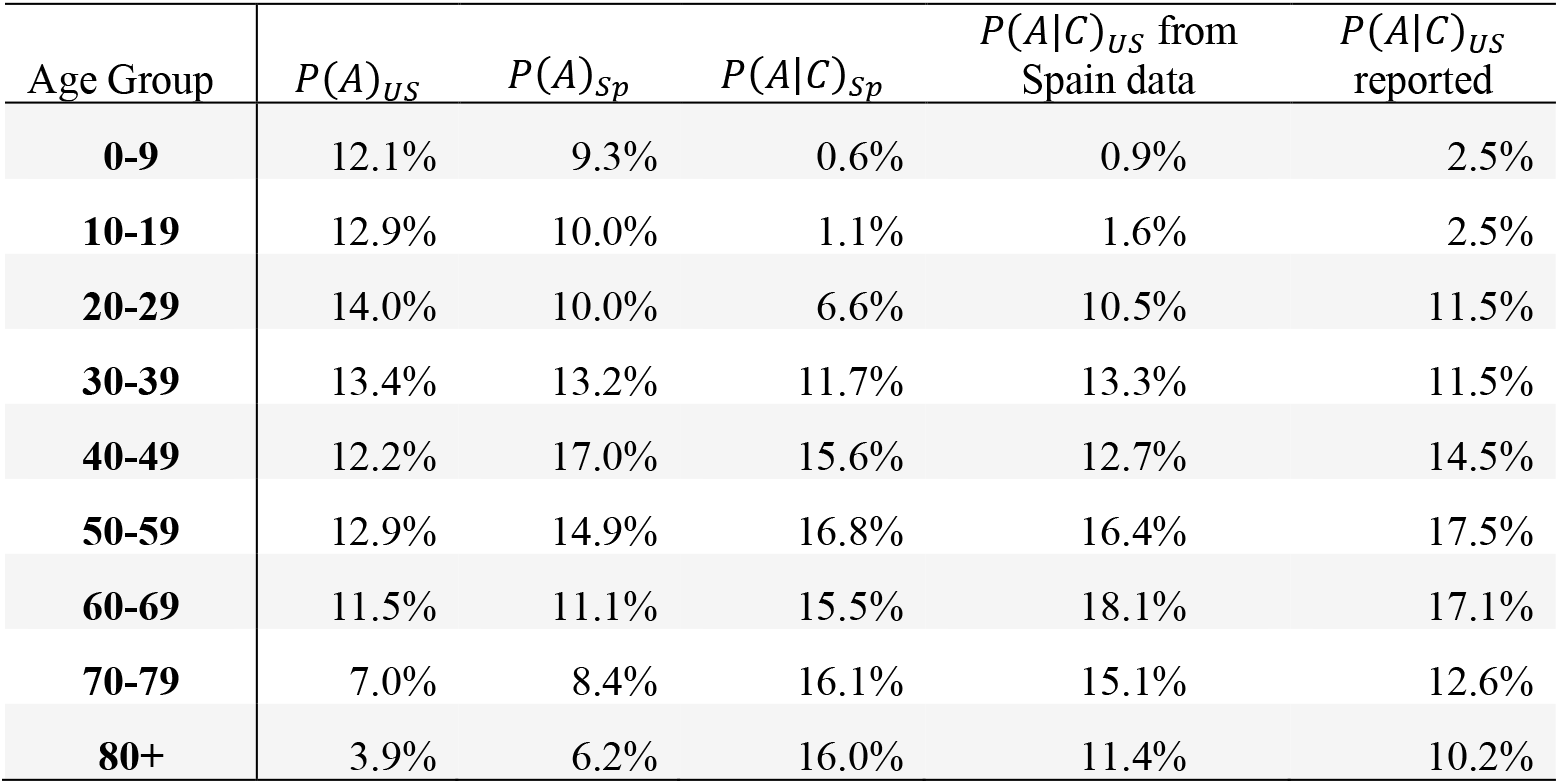
Computing age group probabilities for given cases in (i) Spain, (ii) in the US calculated using Spain data, and (iii) reported by CDC. While the correlation between (i) and (iii) is.88, the correlation between (i) and (ii) is as high as.97.

| Age Group | $P(A)_{US}$ | $P(A)_{Sp}$ | $P(A\vert C)_{Sp}$ | $P(A\vert C)_{US}$ from Spain data | $P(A\vert C)_{US}$ reported |
| --- | --- | --- | --- | --- | --- |
| **0-9** | 12.1% | 9.3% | 0.6% | 0.9% | 2.5% |
| **10-19** | 12.9% | 10.0% | 1.1% | 1.6% | 2.5% |
| **20-29** | 14.0% | 10.0% | 6.6% | 10.5% | 11.5% |
| **30-39** | 13.4% | 13.2% | 11.7% | 13.3% | 11.5% |
| **40-49** | 12.2% | 17.0% | 15.6% | 12.7% | 14.5% |
| **50-59** | 12.9% | 14.9% | 16.8% | 16.4% | 17.5% |
| **60-69** | 11.5% | 11.1% | 15.5% | 18.1% | 17.1% |
| **70-79** | 7.0% | 8.4% | 16.1% | 15.1% | 12.6% |
| **80+** | 3.9% | 6.2% | 16.0% | 11.4% | 10.2% |

We then use country demographics, CDC estimations for the US, and data sets available (shown in Table 5) to compute age-adjusted probabilities and number of cases for each of the events (Susceptible, Case with symptoms, Hospitalization, IUC case, and Deaths). For all countries, we report all numbers per 1 million for easy comparison in Appendix Table 1 Appendix Table 2.

**Table 5.** Datasets used in this study

| Country | South Korea(30) | Spain(35) | US(30) | China(19) | Italy(13) |
| --- | --- | --- | --- | --- | --- |
| Number of cases | 6,284 | 20,471 | 2,449 | 44,669 | ~34,000 |
| Number of hospitalizations | - | 7,699 | - | - | - |
| Number of ICU cases | - | 573 | - | - | - |
| Number of fatalities in the study | 42 | 805 | 44 | 805 | 1,625 |

Studies report different case- and death-related age-breakdowns for a variety of countries. We observed that taking conditional probabilities—based on population age distributions in individual countries—into account, we can help mitigate the variability in the reported results. Figure 2 visually confirms the reduction in inconsistencies by taking age adjustments into account.

**Figure 2.**
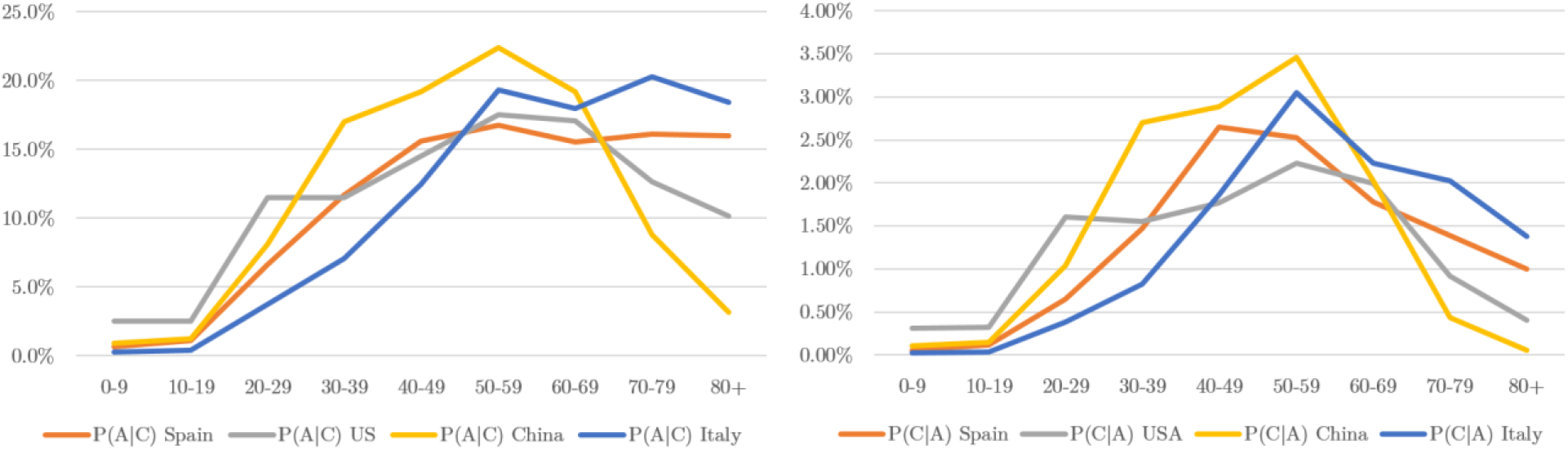
Age breakdown patterns of cases with (left) and without (right) taking country population pyramids (in terms of conditional probabilities) into account.

Figure 3 also highlights the age distribution differences for different events. ICU beds and invasive ventilators are in short supply, and some health systems prioritize younger patients over the older ones in order to increase the chances of survival. While debated from ethical viewpoints, the figure also demonstrates such preferences.

**Figure 3.**
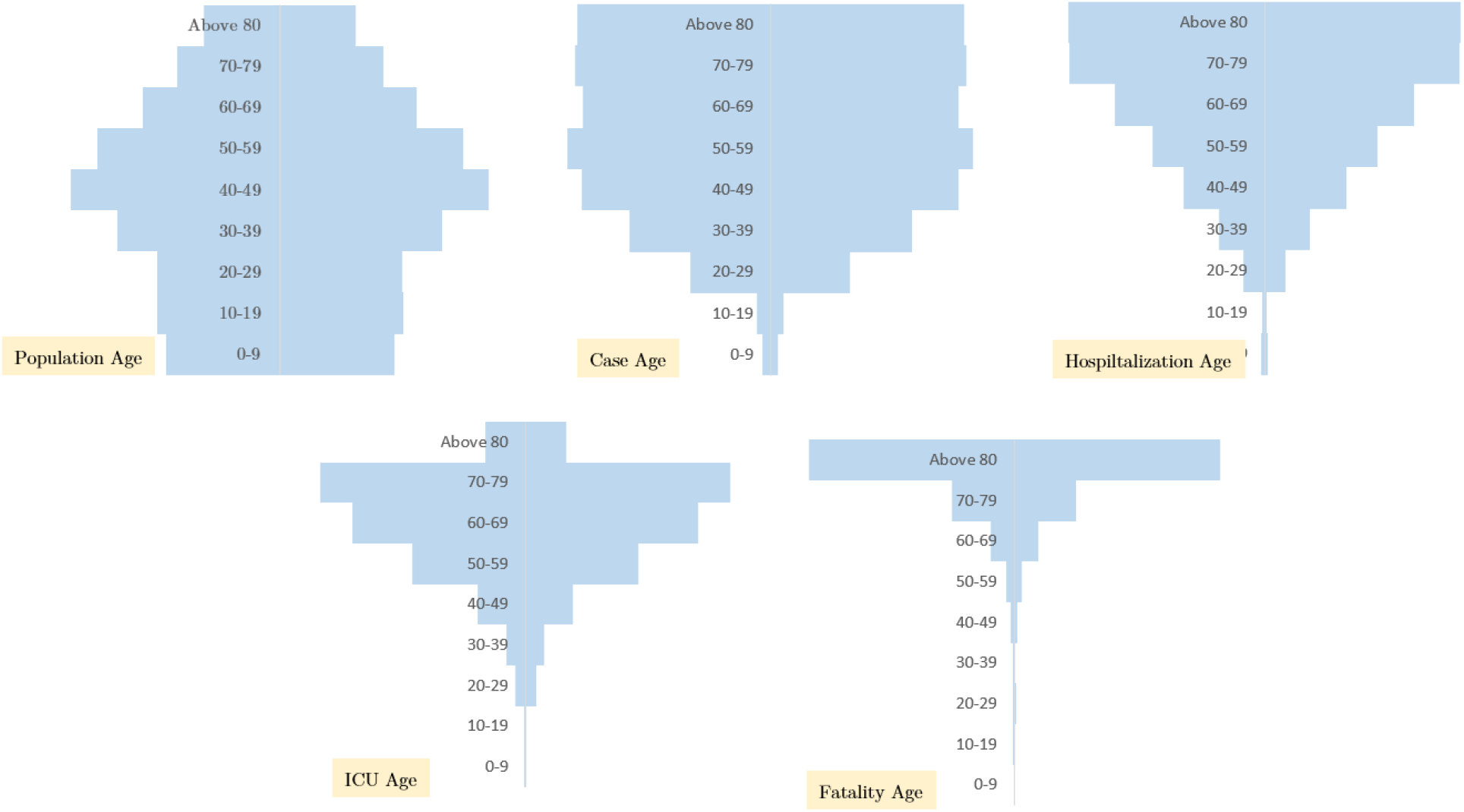
Age pyramids for cases, hospitalization, ICU uses, and fatalities for Spain.

### Assessing Countries’ Reporting Quality

As explained earlier, for each country, we calculated a dissonance ratio (DR) number indicating how well the governments have done in conducting tests and reporting cases, given some age-standardized expected number of cases. The DR estimates (along with other related measures) are reported in detail in Appendix Table 2.

At the time of writing this manuscript, some of the countries were still experiencing the early stages of the breakout. At the end of the COVID-19 spread, the Appendix Table 2 would include similar values in column AARFPM across all countries. As AARFPM values indicate the progress for the breakout, we filtered out the countries with less than 10 AARFPM for better interpretability. This column, therefore, may be interpreted as a crude measure of reliability (hence progress of the spread) for dissonance ratios (i.e., the higher this value, the more evidence we have for the magnitude of the dissonance). Therefore, the table is ordered by AARFPM values rather than dissonance ratios. The correct way of interpreting the results is by comparing the DR values of row-wise nearby countries (with similar AARFPM values). For instance, Finland, Hungary, and Israel have similar AARFPM values (20.1, 20.2, and 20.6, respectively) indicating them being at the same stage of the epidemic. However, their Dissonance Ratios are 0.04, 0.23, and 0.55, respectively. This indicates comparatively better case reporting (including testing) by Israel. At the end of the pandemic, all countries in the table are expected to have roughly the same AARFPM values, making reporting quality of them all comparable to one another through the proposed standardized DR measure.

## Discussion

In this paper, we focus on age-dependent breakdowns of cases, hospitalizations, ICU usages, and fatalities (events) using a range of scenarios. We construct these scenarios by using expert views and existing reports in the literature and based on the US data. We then use conditional probabilities to compute age-standardized breakdowns for the events for all individual countries.

Our results propose a few important implications. First, the results highlight the effect of demographical differences across countries on COVID-19 spread. Figure 4 indicates a comparison between Niger and Japan (as the youngest and oldest populations in the world, respectively). It suggests, provided that everything else remains the same, the death toll difference due to age demographics could be as much as 20 times (47 vs. 1,037 deaths per 1M population according to Appendix Table 1).

**Figure 4.**
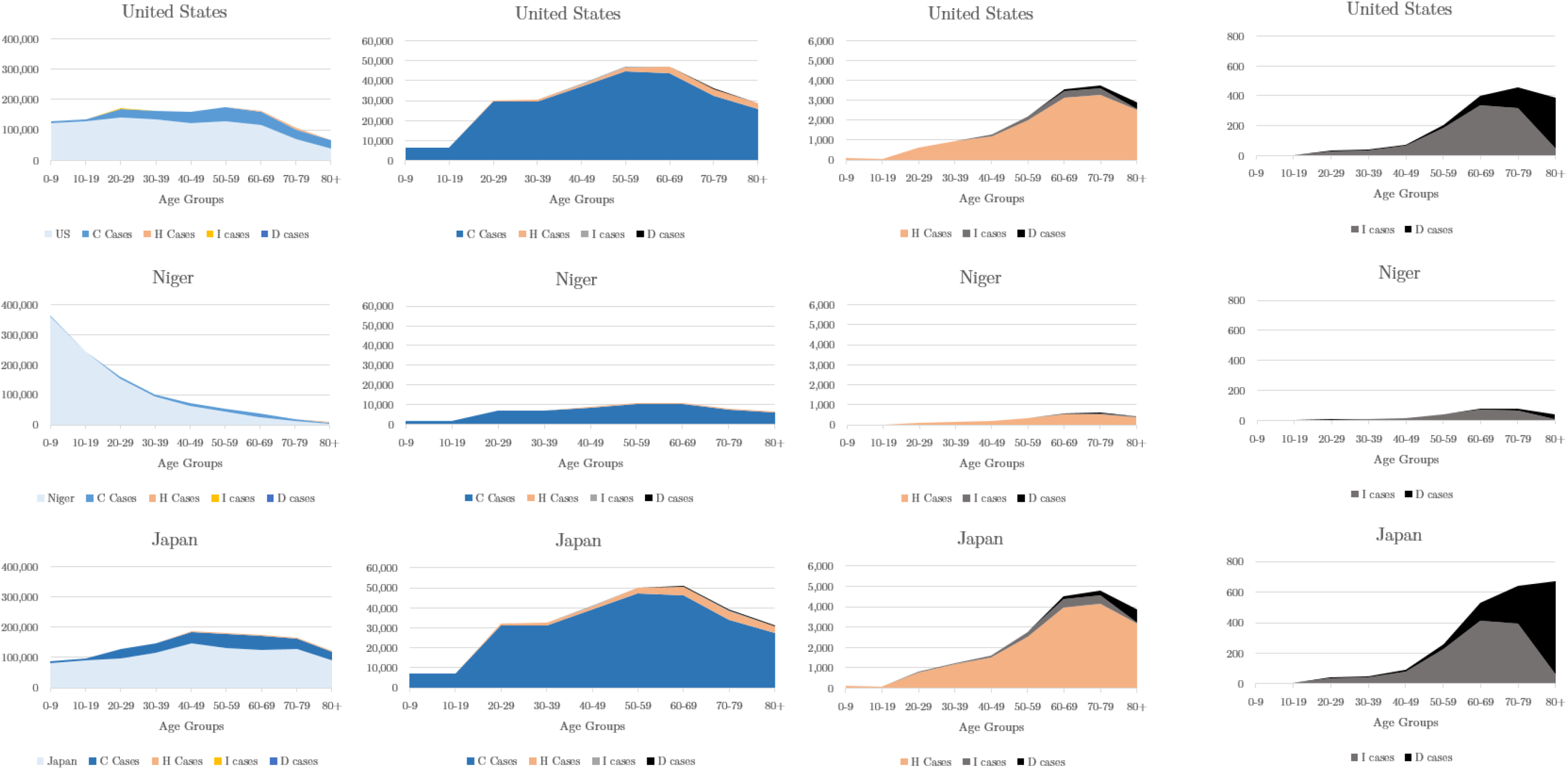
Age-dependent event estimations for the US, the country with the youngest population in the world (Niger), and with the oldest (Japan). y-axis is for scaling purposes based on one of the scenarios per 1M.

Second, our results have the potential to help decision-makers to accommodate age-specific aspects of the spread. Creating different age-based isolation strategies, depending on the age-demographics of individual countries, may be considered. This is essentially important given the limited capacity of health care systems in the affected countries. At this time, one of the major concerns of the governments in the countries affected by COVID-19 is to have reliable estimates of medical requirements to plan ahead proper measures. We argue that providing such estimates based on age groups can give the authorities a clearer picture of what they should expect in different regions of their countries based on the demographic profile of the population in each region. The proposed approach is not exclusively for COVID-19 and may be employed by the decision-makers for estimating capacity requirements in the future epidemic situations.

Also, our study attempts to combine several parameters calculated or taken from different academic papers, reports, or data sources together in creating a range of scenarios. While this approach provides a somewhat holistic view of the phenomenon, it also omits other country-level differences such as social isolation policies, prevention strategies, and the effectiveness of the individual healthcare systems (i.e., our research limitations). Future research may extend the proposed approach by involving such factors (upon the availability of data for them) in the calculation of conditional probabilities. Appendix Table 1 must be interpreted as a comparison tool for different countries’ exposure to the virus.

Additionally, we propose an approach to compare countries’ performance in reporting COVID-19 cases. Clearly, not all countries have the same healthcare infrastructure to conduct enough number of tests to identify infected people. Despite these differences, official reports published by the governments of different countries are being used in a similar manner to study the characteristics of the novel coronavirus, which may cause significant bias to their findings. It is crucial then to provide a means to recognize those countries that perform better in running tests and reporting cases, thereby providing more reliable numbers for studying the pandemic. Appendix Table 2 lays out a base for comparing the reporting performance of the countries by taking into account their age disparities as well as the progress stage of the disease. Using that tool, researchers could have a better understanding of the accuracy of reported numbers by each country by looking into their dissonance ratio number.

## Data Availability

-

## Appendix

**Appendix Table 1.** Scenario-2 based event breakdowns for individual countries

| Country | Population (K) | Cases per 1M | Hosp. per 1M | ICU per 1M | Deaths per 1M |
| --- | --- | --- | --- | --- | --- |
| Portugal | 10,196 | 275,770 | 16,717 | 1,218 | 854 |
| Germany | 83,783 | 275,671 | 16,521 | 1,194 | 852 |
| Italy | 60,461 | 275,463 | 16,903 | 1,223 | 902 |
| Greece | 10,423 | 273,517 | 16,472 | 1,173 | 879 |
| Switzerland | 8,655 | 272,148 | 15,554 | 1,130 | 720 |
| Bulgaria | 6,949 | 271,055 | 16,053 | 1,216 | 706 |
| Japan | 126,475 | 271,042 | 17,430 | 1,256 | 1,037 |
| Hungary | 9,660 | 270,439 | 15,464 | 1,160 | 665 |
| Finland | 5,542 | 270,389 | 16,135 | 1,195 | 784 |
| Netherlands | 17,136 | 269,922 | 15,626 | 1,169 | 705 |
| Spain | 46,733 | 269,495 | 15,673 | 1,117 | 772 |
| Republic of Korea | 51,270 | 269,133 | 14,500 | 1,102 | 556 |
| Romania | 19,236 | 268,786 | 15,330 | 1,133 | 677 |
| Poland | 37,847 | 267,820 | 15,109 | 1,113 | 656 |
| Belgium | 11,589 | 267,496 | 15,314 | 1,104 | 736 |
| Denmark | 5,792 | 266,784 | 15,340 | 1,144 | 694 |
| Czech | 10,708 | 265,466 | 15,251 | 1,153 | 642 |
| Sweden | 10,099 | 264,504 | 15,248 | 1,108 | 729 |
| United Kingdom | 67,886 | 264,035 | 14,909 | 1,078 | 694 |
| Ukraine | 43,734 | 263,634 | 14,445 | 1,065 | 598 |
| Norway | 5,421 | 261,559 | 14,405 | 1,060 | 619 |
| Cuba | 11,326 | 260,576 | 13,965 | 1,038 | 562 |
| Belarus | 9,449 | 256,685 | 13,641 | 1,004 | 548 |
| US | 329,064 | 255,500 | 13,797 | 1,021 | 575 |
| New Zealand | 4,822 | 254,069 | 13,778 | 1,022 | 576 |
| Australia | 25,498 | 253,440 | 13,696 | 993 | 590 |
| Russian Federation | 145,935 | 253,217 | 13,567 | 997 | 551 |
| Uruguay | 3,474 | 244,333 | 12,945 | 912 | 586 |
| Ireland | 4,938 | 242,002 | 12,670 | 941 | 493 |
| Chile | 19,116 | 234,534 | 11,579 | 853 | 429 |
| China | 1,439,324 | 227,809 | 10,805 | 875 | 326 |
| Argentina | 45,197 | 213,004 | 10,468 | 759 | 400 |
| Israel | 8,655 | 211,875 | 10,745 | 771 | 438 |
| Sri Lanka | 21,413 | 211,258 | 10,060 | 822 | 304 |
| Brazil | 212,560 | 207,735 | 9,592 | 722 | 318 |
| Colombia | 50,883 | 198,163 | 9,060 | 679 | 301 |
| **World** | **<u>7,794,799</u>** | **<u>197,313</u>** | **<u>9,131</u>** | **<u>688</u>** | **<u>304</u>** |
| Tunisia | 11,819 | 196,661 | 8,884 | 680 | 281 |
| Turkey | 84,339 | 195,387 | 8,928 | 677 | 290 |
| Peru | 32,972 | 191,468 | 8,717 | 649 | 290 |
| Viet Nam | 97,338 | 190,509 | 8,391 | 608 | 284 |
| Kazakhstan | 18,776 | 181,352 | 8,042 | 604 | 258 |
| Venezuela (Bolivarian Republic of) | 28,437 | 180,853 | 8,072 | 628 | 247 |
| Mexico | 128,933 | 177,978 | 7,887 | 589 | 256 |
| Dominica | 10,847 | 174,998 | 7,815 | 574 | 263 |
| Azerbaijan | 10,139 | 173,391 | 7,261 | 557 | 214 |
| Ecuador | 17,643 | 173,165 | 7,726 | 571 | 257 |
| Morocco | 36,910 | 172,690 | 7,503 | 604 | 214 |
| Malaysia | 32,365 | 168,392 | 7,211 | 575 | 205 |
| Bolivia | 11,674 | 163,184 | 7,436 | 533 | 267 |
| Iran (Islamic Republic of) | 83,993 | 161,603 | 6,787 | 541 | 189 |
| Algeria | 43,852 | 159,997 | 6,934 | 521 | 217 |
| Paraguay | 7,132 | 155,116 | 6,797 | 516 | 213 |
| India | 1,380,004 | 154,638 | 6,475 | 529 | 176 |
| Indonesia | 273,523 | 154,124 | 6,343 | 531 | 165 |
| Bangladesh | 164,690 | 142,614 | 5,915 | 448 | 177 |
| Philippines | 109,581 | 136,985 | 5,625 | 457 | 152 |
| Nepal | 29,138 | 133,698 | 5,539 | 465 | 145 |
| South Africa | 59,308 | 132,704 | 5,360 | 455 | 135 |
| Turkmenistan | 6,031 | 129,106 | 5,120 | 398 | 143 |
| Egypt | 102,335 | 129,065 | 5,306 | 440 | 140 |
| Uzbekistan | 33,470 | 128,528 | 5,038 | 401 | 135 |
| Syrian Arab Republic | 17,500 | 125,473 | 5,083 | 400 | 141 |
| Pakistan | 220,892 | 112,174 | 4,521 | 370 | 119 |
| Jordan | 10,205 | 108,890 | 4,301 | 345 | 114 |
| Saudi Arabia | 34,815 | 102,129 | 3,777 | 317 | 91 |
| Sudan | 43,849 | 93,224 | 3,681 | 308 | 93 |
| Ethiopia | 114,964 | 88,752 | 3,545 | 291 | 93 |
| Iraq | 40,223 | 88,523 | 3,410 | 280 | 87 |
| Iraq | 40,223 | 88,523 | 3,410 | 280 | 87 |
| South Sudan | 11,195 | 85,682 | 3,366 | 285 | 84 |
| State of Palestine | 5,101 | 85,658 | 3,307 | 278 | 82 |
| Madagascar | 27,692 | 82,226 | 3,138 | 265 | 77 |
| Ghana | 31,073 | 79,587 | 2,934 | 280 | 63 |
| Senegal | 16,745 | 77,735 | 2,989 | 261 | 71 |
| Democratic Republic of the Congo | 89,561 | 76,412 | 2,970 | 255 | 72 |
| Yemen | 29,825 | 74,766 | 2,840 | 250 | 67 |
| Mozambique | 31,255 | 71,548 | 2,752 | 239 | 66 |
| Somalia | 15,893 | 70,564 | 2,714 | 242 | 63 |
| Côte d'Ivoire | 26,378 | 68,960 | 2,558 | 243 | 55 |
| Cameroon | 26,545 | 68,088 | 2,540 | 232 | 57 |
| United Republic of Tanzania | 59,734 | 66,731 | 2,484 | 229 | 55 |
| Afghanistan | 38,928 | 66,159 | 2,469 | 227 | 55 |
| Kenya | 53,771 | 66,064 | 2,407 | 219 | 53 |
| Nigeria | 206,139 | 60,712 | 2,193 | 228 | 43 |
| Burkina Faso | 20,903 | 60,006 | 2,200 | 209 | 47 |
| Mali | 20,249 | 59,879 | 2,230 | 207 | 49 |
| Niger | 24,207 | 59,262 | 2,215 | 215 | 47 |

**Appendix Table 2.** Measuring the gap between expected case reporting vs. actual case reporting

| Country | Popul. (K) | ACFR | RIPF | EIPF | DR | AARFPM | CF | DACC | AAHC | AAICUC | AAFC |
| --- | --- | --- | --- | --- | --- | --- | --- | --- | --- | --- | --- |
| Belgium | 11,589 | 17.4% | 5.73 | 161 | 0.04 | 310.3 | 8,707 | 49,906 | 50.4 | 3.63 | 2.42 |
| United Kingdom | 67,886 | 17.2% | 5.82 | 157 | 0.04 | 206.9 | 32,065 | 186,599 | 49.0 | 3.55 | 2.28 |
| Italy | 60,461 | 14.6% | 6.86 | 178 | 0.04 | 171.3 | 30,739 | 210,717 | 55.6 | 4.02 | 2.97 |
| Sweden | 10,099 | 14.6% | 6.85 | 161 | 0.04 | 134.4 | 3,256 | 22,317 | 50.2 | 3.64 | 2.40 |
| Hungary | 9,660 | 14.0% | 7.14 | 163 | 0.04 | 20.1 | 425 | 3,035 | 50.9 | 3.82 | 2.19 |
| Netherlands | 17,136 | 13.4% | 7.44 | 165 | 0.05 | 137.2 | 5,456 | 40,571 | 51.4 | 3.85 | 2.32 |
| Spain | 46,733 | 12.3% | 8.13 | 165 | 0.05 | 225.3 | 26,744 | 217,466 | 51.6 | 3.67 | 2.54 |
| Mexico | 128,933 | 15.2% | 6.57 | 85 | 0.08 | 32.9 | 3,573 | 23,471 | 25.9 | 1.94 | 0.84 |
| Romania | 19,236 | 7.4% | 13.54 | 162 | 0.08 | 22.6 | 972 | 13,163 | 50.4 | 3.73 | 2.23 |
| Brazil | 212,560 | 11.4% | 8.78 | 101 | 0.09 | 51.8 | 11,519 | 101,147 | 31.6 | 2.38 | 1.05 |
| US | 329,064 | 7.0% | 14.35 | 145 | 0.10 | 129.6 | 80,684 | 1,158,041 | 45.4 | 3.36 | 1.89 |
| Ireland | 4,938 | 6.8% | 14.66 | 134 | 0.11 | 183.1 | 1,467 | 21,506 | 41.7 | 3.10 | 1.62 |
| Denmark | 5,792 | 5.6% | 17.87 | 162 | 0.11 | 40.3 | 533 | 9,523 | 50.5 | 3.76 | 2.28 |
| Finland | 5,542 | 5.2% | 19.39 | 170 | 0.11 | 18.9 | 271 | 5,254 | 53.1 | 3.93 | 2.58 |
| Switzerland | 8,655 | 5.2% | 19.34 | 164 | 0.12 | 75.2 | 1,542 | 29,822 | 51.2 | 3.72 | 2.37 |
| Algeria | 43,852 | 11.3% | 8.82 | 72 | 0.12 | 16.2 | 507 | 4,474 | 22.8 | 1.71 | 0.71 |
| Germany | 83,783 | 4.6% | 21.66 | 174 | 0.12 | 32.0 | 7,533 | 163,175 | 54.3 | 3.93 | 2.80 |
| **World** | **7,794,799** | **8.2%** | **12.13** | **96** | **0.13** | **36.6** | **285,760** | **3,465,608** | **30.0** | **2.26** | **1.00** |
| Portugal | 10,196 | 4.5% | 22.31 | 176 | 0.13 | 39.9 | 1,144 | 25,524 | 55.0 | 4.01 | 2.81 |
| Czech | 10,708 | 3.6% | 27.59 | 161 | 0.17 | 12.4 | 282 | 7,781 | 50.2 | 3.79 | 2.11 |
| Bolivia | 11,674 | 7.7% | 13.07 | 76 | 0.17 | 11.9 | 122 | 1,594 | 24.5 | 1.75 | 0.88 |
| Ecuador | 17,643 | 7.3% | 13.77 | 77 | 0.18 | 143.8 | 2,145 | 29,538 | 25.4 | 1.88 | 0.85 |
| Iran | 83,993 | 6.9% | 14.57 | 78 | 0.19 | 128.0 | 6,685 | 97,424 | 22.3 | 1.78 | 0.62 |
| Philippines | 109,581 | 7.9% | 12.70 | 62 | 0.20 | 13.2 | 726 | 9,223 | 18.5 | 1.50 | 0.50 |
| Egypt | 102,335 | 8.2% | 12.13 | 54 | 0.22 | 11.3 | 533 | 6,465 | 17.5 | 1.45 | 0.46 |
| Norway | 5,421 | 2.9% | 34.86 | 152 | 0.23 | 20.2 | 224 | 7,809 | 47.4 | 3.49 | 2.04 |
| Dominican Republic | 10,847 | 4.9% | 20.24 | 83 | 0.24 | 41.8 | 393 | 7,954 | 25.7 | 1.89 | 0.87 |
| Peru | 32,972 | 4.3% | 23.42 | 94 | 0.25 | 62.3 | 1,961 | 45,928 | 28.7 | 2.13 | 0.95 |
| Turkey | 84,339 | 3.0% | 32.82 | 94 | 0.35 | 47.7 | 3,841 | 126,045 | 29.4 | 2.23 | 0.95 |
| Somalia | 15,893 | 7.2% | 13.88 | 29 | 0.48 | 15.7 | 52 | 722 | 8.9 | 0.80 | 0.21 |
| Chile | 19,116 | 1.6% | 60.88 | 122 | 0.50 | 11.9 | 323 | 19,663 | 38.1 | 2.81 | 1.41 |
| Israel | 8,655 | 1.6% | 62.82 | 113 | 0.55 | 20.6 | 258 | 16,208 | 35.3 | 2.54 | 1.44 |
| Burkina Faso | 20,903 | 7.6% | 13.24 | 23 | 0.57 | 15.4 | 50 | 662 | 7.2 | 0.69 | 0.15 |
| Cameroon | 26,545 | 6.0% | 16.62 | 27 | 0.62 | 25.1 | 125 | 2,077 | 8.4 | 0.76 | 0.19 |
| Mali | 20,249 | 6.9% | 14.44 | 23 | 0.62 | 11.9 | 39 | 563 | 7.3 | 0.68 | 0.16 |
| Niger | 24,207 | 6.1% | 16.30 | 24 | 0.69 | 12.2 | 46 | 750 | 7.3 | 0.71 | 0.15 |
| Afghanistan | 38,928 | 4.5% | 22.16 | 26 | 0.85 | 17.3 | 122 | 2,704 | 8.1 | 0.75 | 0.18 |
| Saudi Arabia | 34,815 | 0.9% | 105.93 | 45 | 2.34 | 24.4 | 255 | 27,011 | 12.4 | 1.04 | 0.30 |

## Notes

### Competing Interest Statement

The authors have declared no competing interest.

### Funding Statement

The research received no funding from any sources.

